# Evaluating the life-extending potential and safety profile of rapamycin: a Mendelian Randomization study of the mTOR pathway

**DOI:** 10.1101/2023.10.02.23296427

**Authors:** Maria K Sobczyk, Tom R Gaunt

## Abstract

**Objective:** The mechanistic target of rapamycin (mTOR) pathway plays an integral role in cellular metabolism, growth, and aging. While rapamycin and its analogs inhibit the mTOR pathway, extending lifespan in various organisms, the long-term safety and efficacy of these compounds in humans remain understudied.

**Methods:** Utilizing two mTOR expression QTL instruments derived from the eQTLgen and MetaBrain studies, we sought to explore the potential causal relationship between mTOR expression inhibition in blood and brain cortex (mimicking chronic rapamycin use), and its effects on longevity, cardiometabolic disease, prostate cancer and anthropometric risk factors. Subsequently, we extended the selection of instruments to 47 other members of the mTOR pathway. To complement this Mendelian randomization (MR) evidence, we conducted genetic colocalisation and sampling-based enrichment testing.

**Results:** Our findings suggest that genetically proxied mTOR inhibition may increase the odds of attaining top 1% longest lifespan in the population (OR=1.24, OR_95%CI_=1-1.53, p-value=0.048). Moreover, mTOR inhibition significantly reduced body mass index (BMI), basal metabolic rate (BMR), height, and age at menopause, while increasing bone mineral density. Interestingly, there was generally little evidence linking mTOR inhibition to cardiovascular disease incidence, with the exception of weak evidence for a protective effect against heart failure (OR=0.94, OR_95%CI_=0.89-0.99, p-value=0.039). Chronic mTOR inhibition did not causally affect prostate cancer incidence but increased the risk of developing type 2 diabetes. A higher-than-expected (p-value = 0.05) number of genes in the mTOR pathway were causally associated with BMR.

**Conclusions:** This study highlights the potential lifespan-extending effects of mTOR inhibition and its significant influence on metabolic risk factors and disease. Members of the mTOR complex, especially mTORC1, play a disproportionate role in influencing BMR and BMI, which provides valuable insight for potential therapeutic target development.

## Introduction

The mechanistic target of rapamycin (mTOR) pathway plays a crucial role in regulating cellular metabolism, growth, and aging^1^. It is centred on the mechanistic target of rapamycin (mTOR), a protein kinase that integrates various environmental and intracellular signals. The pathway is composed of two protein complexes, mTOR complex 1 (mTORC1) and mTOR complex 2 (mTORC2), which have different substrates and functions but are inter-linked^2^.

mTORC1 is activated by growth factors, nutrients, and energy status, and regulates protein synthesis, autophagy, and metabolism^3^. mTORC1 is directly activated by the PI3K/AKT pathway through inhibition of the TSC1/TSC2 complex, which acts as a negative regulator of Rheb GTPase which in turn stimulates mTORC1. mTORC1’s main substrates include S6 kinase 1 (S6K1) and eukaryotic translation initiation factor 4E-binding protein (4E-BP), which control mRNA translation and protein synthesis. mTORC1 also promotes lipid synthesis and inhibits autophagy through its phosphorylation of the Unc-51-like kinase 1 (ULK1).

mTORC2 is activated by insulin, IGF-1, and leptin, and regulates cell survival, cytoskeletal organization, and glucose metabolism^4^. mTORC2 phosphorylates AKT at S473, which enhances its activity and leads to feedback activation of mTORC1 via PRAS40 (AKT1S1) phosphorylation. mTORC2’s main substrate is serine/threonine-protein kinase 1 (SGK1), which controls cell survival and proliferation. mTORC2 also modulates actin cytoskeleton organization and motility through its regulation of protein kinase Cα(PKCα) and Rho GTPases.

While mTORC1 and mTORC2 share many common components, such as Deptor and mLST8, they differ in regulatory mechanisms and downstream signalling pathways. Dysregulation of either complex has been implicated in various diseases, including cancer, type 2 diabetes, and aging in general^1^.

mTOR is a drug target for rapamycin (sirolimus), a macrolide agent first isolated from *Streptomyces* bacteria on Rapa Nui (Easter Island)^5^. In addition, several analogs of rapamycin (rapalogs), such as everolimus, are in clinical use due to their improved pharmacokinetic properties^6,7^. Inhibition of mTORC1 with rapamycin or its analogs has been shown to extend lifespan and delay age-related diseases across many model organisms, from yeast to mammals^8^. In mice, rapamycin increased mean lifespan by 9-14% when started at middle age, which is similar in magnitude to the effect observed in S6K1 loss-of-function mutants and Mtor+/–Mlst8+/– heterozygotes^6,9^. Rapamycin can also delay age-related disorders, such as cancer, cardiac dysfunction, cognitive decline and immune senescence^10^. While it partially mimics the effect of calorie restriction, the precise mechanism of lifespan extension by rapamycin is not fully understood but may involve modulation of nutrient sensing pathways, autophagy induction, inflammation reduction, stem cell renewal, inhibition of protein translation and others^6,11^.

Low-dose and long-term unauthorised rapamycin use has been gaining interest among anti-aging “biohackers” as a longevity-promoting intervention in humans and dogs^12–15^. Randomized clinical trials are ongoing, with some early results indicating potential benefits in immune function in phase 2 trials^16,17^. PEARL (Participatory Evaluation of Aging with Rapamycin for Longevity), a randomized, placebo-controlled phase 4 trial aiming to enrol 1000 healthy older adults will assess the safety and efficacy of rapamycin in improving visceral fat content, bone mass density and lean body mass^18^.

However, long-term safety and efficacy data is still lacking and will continue to be so given the short duration of intervention in clinical trials (e.g. 24 weeks for PEARL). Collected endpoints measured in clinical trials will reflect short-term changes in cardiometabolic and immune health which may not be adequate proxies for extended longevity. Moreover, long-term rapamycin treatment can have undesirable side effects such as immunosuppression, insulin resistance leading to type 2 diabetes^19^, and increased risk of prostate cancer^20^ due to off-target inhibition of mTORC2^9^. Study designs of current clinical trials will make it difficult to uncover such side-effects of chronic rapamycin use (as potentially experienced when pursuing longevity).

To fill this gap, in this study we aim to investigate the effect of mTOR pathway inhibition on longevity and adverse side effects in humans using a genetic approach called Mendelian randomization (MR). This method uses genetic variants (typically single nucleotide polymorphisms, SNPs) that are known to be associated with the risk factor of interest but not directly related to the outcome, as instrumental variables. The genetic variants are randomly assigned to individuals at conception and are therefore not subject to the confounding factors that may affect observational studies^21,22^. Once the instrumental variables are identified, they can be used to approximate the causal effect of mTOR inhibition by assessing their association with the outcome of interest in large-scale genome-wide association study (GWAS) data sets. As MR estimates are closer to representing lifetime rather than short-term effects of a given intervention^23^, they can reveal the potential benefits and risks associated with chronic rapamycin use.

The MR approach has recently been extended to employ protein quantitative trait loci (pQTLs) – variants associated with protein levels, and expression QTL (eQTLs) – variants associated with gene transcript levels, which can serve as useful proxies for the impact of modulating such targets by drugs^24^. Three main goals of drug target perturbation MR concern: validation of hypothesized drug effects^25–27^, prediction of adverse effects^28,29^ and repurposing of existing therapeutics^27,30,31^; here we focus on the last two uses.

Previous research employed genetic variants that modulate the expression of mTORC1’s downstream targets regulating protein translation to study their effect on type 2 diabetes, rheumatic fever, Alzheimer’s disease, Parkinson’s disease, and cataract formation^32–36^. These studies utilised moderately associated (p-value < 5 × 10^-6^) and independent (linkage disequilibrium r^2^ < 0.05) variants associated with protein expression in blood plasma. In this current study, we extend that selection to include the mTOR protein, both mTORC1 and mTORC2 along with their select upstream regulators and downstream targets. Using stringently selected plasma pQTL and eQTL genetic instruments from blood and brain cortex, we attempt to model rapamycin’s inhibitory effect on longevity, anthropometric traits and disease.

## Materials and Methods

### Genetic exposure data sources

We used KEGG^37^ and a number of reviews^1–4^ on mTOR pathway biology to select the 48 gene exposures used in the analysis, which correspond to the specific proximal regulators and effectors of mTORC1 and mTORC2 which are associated with *cis-*eQTL variants or pQTL variants in the following studies: MetaBrain cortex eQTL (European)^38^ and blood plasma eQTLs from eQTLgen^39^ and European pQTLs from ARIC^40^, deCODE^41^, FENLAND^42^, UK Biobank (UKBB)^43^ (**Supplementary Table 1-2, Figure 1**). We used those tissues due to high expression of mTOR in the brain and blood^44^, high metabolic activity of the brain^45^ and blood expression reflecting overall physiological state of the organism^46^. In addition, previous research has shown that eQTL effects tend to be shared among brain and non-brain tissues^47^, so our study includes a single representative from each group based on availability and sample size (and therefore statistical power).

**Figure 1.**
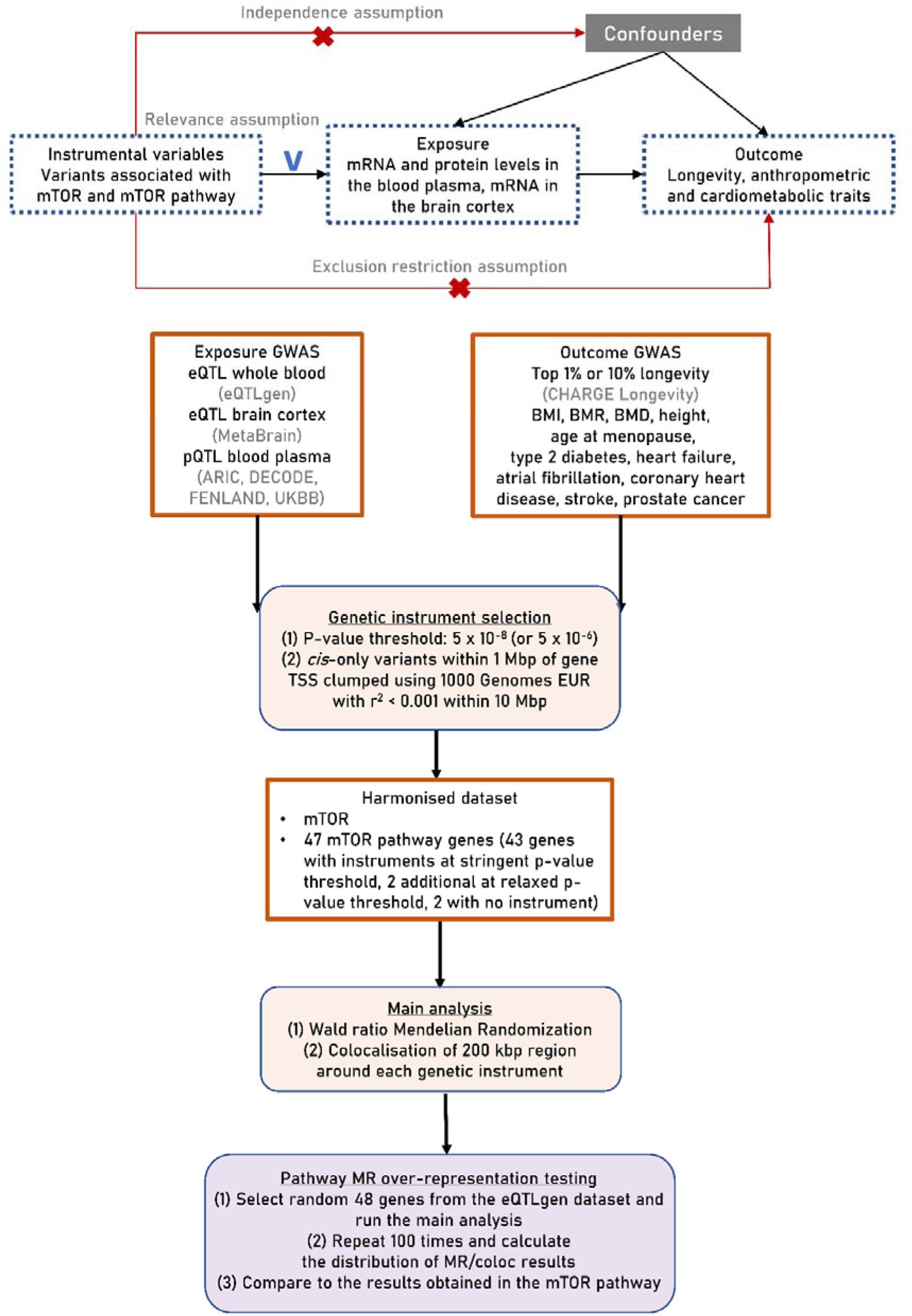
Key assumptions of Mendelian Randomization and study overview. *BMI* - body mass index, *BMR* - basal metabolic rate, *BMD* - bone mineral density, *eQTL* - expression quantitative trait locus, *pQTL* - protein quantitative trait locus.

### Genetic outcome data sources

We used the CHARGE Longevity GWAS meta-analysis for top 90^th^ percentile and 99^th^ percentile^48^ lifespan as our longevity outcome for mTOR inhibition. This is the biggest GWAS to date directly looking at individuals identified to have the longest lifespan in their birth cohort. ‘Cases’ were participants who survived beyond either the 90th or 99th percentile of their birth cohort’s lifespan, determined using demographic data from the relevant country. On the other hand, ‘controls’ were those who either died by or were last checked up by the age at the 60th percentile of their birth cohort’s lifespan. Various resources such as the Human Mortality Database, the US Social Security Administration, and national registries provided the cohort life tables. For instance, for the US 1920 birth cohort, men reaching the 60th, 90th, and 99th percentiles lived 75, 89, and 98 years respectively, while women lived 83, 94, and 102 years, respectively. When cohort life tables listed birth cohort by decade, yearly birth cohort survival percentiles were estimated using linear model predictions.

To narrow down pleiotropic and off-target effects of mTOR inhibition, we mined EpigraphDB^49^ (https://epigraphdb.org/xqtl) for previous MR results showing significant association of eQTL expression in the blood in eQTLgen with a number of anthropometric and disease outcomes detected (height, weight, body mass index (BMI), age at menopause, heel/total bone mineral density, type 2 diabetes: **Supplementary Table 2**). We were then interested to replicate those results using our chosen exposures, and also extend the analysis to a wide network of mTORC1/2 key players. Given mTOR’s role as a grand conductor of metabolism^4,50,51^, we also evaluate basal metabolic rate as an outcome (**Supplementary Table 4**). We also considered if potentially life-extending properties of rapamycin were mediated through overall improved cardiac function, as previously observed in mice and currently trialled in humans (ClinicalTrials.gov: NCT04996719)^52–54^. Thus, we included common cardiovascular disease phenotypes: coronary heart disease, myocardial infarction, heart failure, atrial fibrillation and stroke, and additional 14 cardiac function traits (listed in Supplementary Table 4). Kidney transplant recipients receiving rapamycin were reported to be at an elevated risk of prostate cancer in an observational study and in a meta-analysis of randomized controlled trials (RCT)^20^. Previously, mTOR inhibition has been targeted as an intervention for a related outcome (prostate cancer progression^55^) which is currently on a small scale trial in humans^56^. Therefore, we also consider prostate cancer incidence outcome. Summary statistics for these outcome GWAS were extracted from OpenGWAS^57^.

### Two-sample Mendelian Randomization analyses

We selected our genetic instruments using a *stringent*, genome-wide p-value threshold of 5 x 10^-8^. To see if a relaxation of the p-value threshold increases the number of genes with a valid genetic instrument, we subsequently also tested a *relaxed* p-value threshold of 5 x 10^-6^ which resulted in addition of eQTL instruments for 2 genes (*NPRL2*, *RHOA*) and pQTL instruments for 1 gene (*AKT1*). We chose to include these additional instruments in our analyses, but for all the other genetic instruments we maintain the *stringent* p-value threshold. We used only *cis-* acting variants located in close proximity to the gene (within 1 Mbp of transcription start site, TSS) as such instruments are less likely to be horizontally pleiotropic, ie. affecting the outcome through expression of multiple genes^58^ and violating the exclusion restriction MR assumption. Furthermore, for pQTL instruments, we removed SNPs (or correlated proxies at r^2^ > 0.8 in 1000 Genomes European) with potentially epitope-altering mutations (“stop_gained”, “stop_lost”, “frameshift_variant”, “start_lost”, “inframe_insertion”, “inframe_deletion”, “missense_variant”, “protein_altering_variant”) identified by Ensembl Variant Effect Predictor (VEP)^59^, as done previously^27^.

To prevent multiple instruments from detecting the same causal effect, we performed clumping of the SNPs to ensure that the linkage disequilibrium (LD) - quantified by r^2^ - was below 0.001 within a 10 Mbp range in the 1000 Genomes European panel^60^. This procedure was conducted using the ld_clump function from the ieugwasr R package (https://mrcieu.github.io/ieugwasr), invoking plink version 1.943^61^. Associations between the genetic variants, exposure and outcome trait were collected and harmonized using the default settings in the TwoSampleMR^62^ package. In that step, we removed all the palindromic SNPs which could not be reliably aligned (minor allele frequency, MAF > 0.42) relative to the same strand and SNPs absent from the outcome dataset. If a given instrument was missing in the outcome data, we tried to establish a proxy with high genetic correlation (r^2^ > 0.8). Finally, we evaluated the F-statistics and R^2^ to identify weak instrument bias.

Two sample MR (Wald ratio for single-SNP instruments) was carried out using default settings in the TwoSampleMR^62^ R package. We used Steiger filtering implemented in the package to assess correct directionality of our MR results.

### Colocalisation evidence

We used the Bayesian colocalisation method coloc^63^ to further establish if significant MR effects observed were likely causal or confounded by LD patterns^64^. We used a 200 kb window around each instrument SNP to define the colocalisation region and run coloc using extracted exposure and outcome summary statistics if at least 50 variants were found in the region. We were interested to identify cases where posterior probability of colocalization (hypothesis 4, H4 - both traits are associated and share a single causal variant) was higher than the other scenarios (> 0.6).

It can be inferred that the power for colocalisation is low if both H3 (both traits are associated, but with different causal variants) and H4 show low posterior probabilities^64^. This conclusion is based on the fact that colocalization was only utilized for cases with a strong association in the exposure dataset (i.e., p-value < 5 x 10^-8^) and supported by MR evidence of a cause-effect relationship. Under this scenario, only H3 or H4 hypotheses are viable, so if the highest posterior probability points to H1 (only trait 1 has a genetic association in the region), it indicates that the outcome dataset may not have enough power for colocalization. Consequently, for significant (p-value < 0.05) MR results which met this criteria, we calculated alternative posterior probability (PP) H4 (referred to as altH4), as follows: PP.H4 / (PP.H3 + PP.H4)^64^ as only these two hypotheses are plausible in this case.

### Random gene set simulation

To see if a high number of MR results with at least marginal evidence (p-value < 0.05) observed for certain outcomes in the mTOR pathway is likely to be a biologically meaningful overrepresentation, we sampled a random 48 gene set with genetic instruments at the *stringent* p-value threshold available in the eQTLgen blood eQTL resource, which contains the highest number of genes with *cis-* instruments overall (15,698). We then ran the MR/coloc analysis described above for 100 randomly drawn gene sets and calculated the empirical distributions of the following statistics: number of significant MR results (p-value < 0.05) and number of coloc results where the (alternative) posterior probability (PP) of hypothesis 4 (H4) > 0.6.

## Results

### Chronic inhibition of mTOR expression may extend longevity

We used single-SNP expression QTL instruments from the blood (eQTLgen study) and brain cortex (MetaBrain study) to investigate the effect of reduced mTOR expression on longevity, and in this way genetically proxy the effect of chronic rapamycin use. Both instruments are strong with F-stats of 757 and 105, respectively, which explain 2.3% and 1.6% of variance in the gene expression, accordingly (**Supplementary Table 5**). Our two-sample MR analysis (**Figure 2**) revealed limited statistical evidence for the effect of *mTOR* gene expression inhibition on being ranked in the 90^th^ percentile with the highest longevity (MetaBrain: OR=1.06, OR_95%CI_=0.93-1.22, p-value=0.38; eQTLgen: OR=1.03, OR_95%CI_=0.91-1.15, p-value=0.66, **Supplementary Table 6**). However, for the top 99^th^ percentile, there was stronger evidence for a positive effect of *mTOR* inhibition on longevity, with odds-ratio of 1.17 based on blood eQTL instrument (OR_95%CI_=0.98-1.41, p-value=0.086) and 1.24 based on the brain eQTL (OR_95%CI_=1-1.53, p-value=0.048). Using Steiger filtering, we found evidence that our interpretation of the direction of causal relationship was correct (**Supplementary Table 7**). Colocalisation analysis (**Supplementary Table 8**) suggested that hypothesis 1 (only trait 1 has a genetic association in the region) had the most support. Since we already found some effect of *mTOR* expression on longevity, we surmised that only hypotheses 3 and 4, involving both traits having an association but with a different or the same variant, respectively, are viable. In that case, the ratio of posterior probability of hypothesis 4 (true colocalisation) to the sum of posterior probabilities of hypotheses 3 and 4, labelled “altH4” here, was previously used^64^. For *mTOR*, we found that hypothesis 4 was supported over hypothesis 3 as altH4 probability rose to 0.75 for the eQTLgen result and 0.79 for MetaBrain.

**Figure 2.**
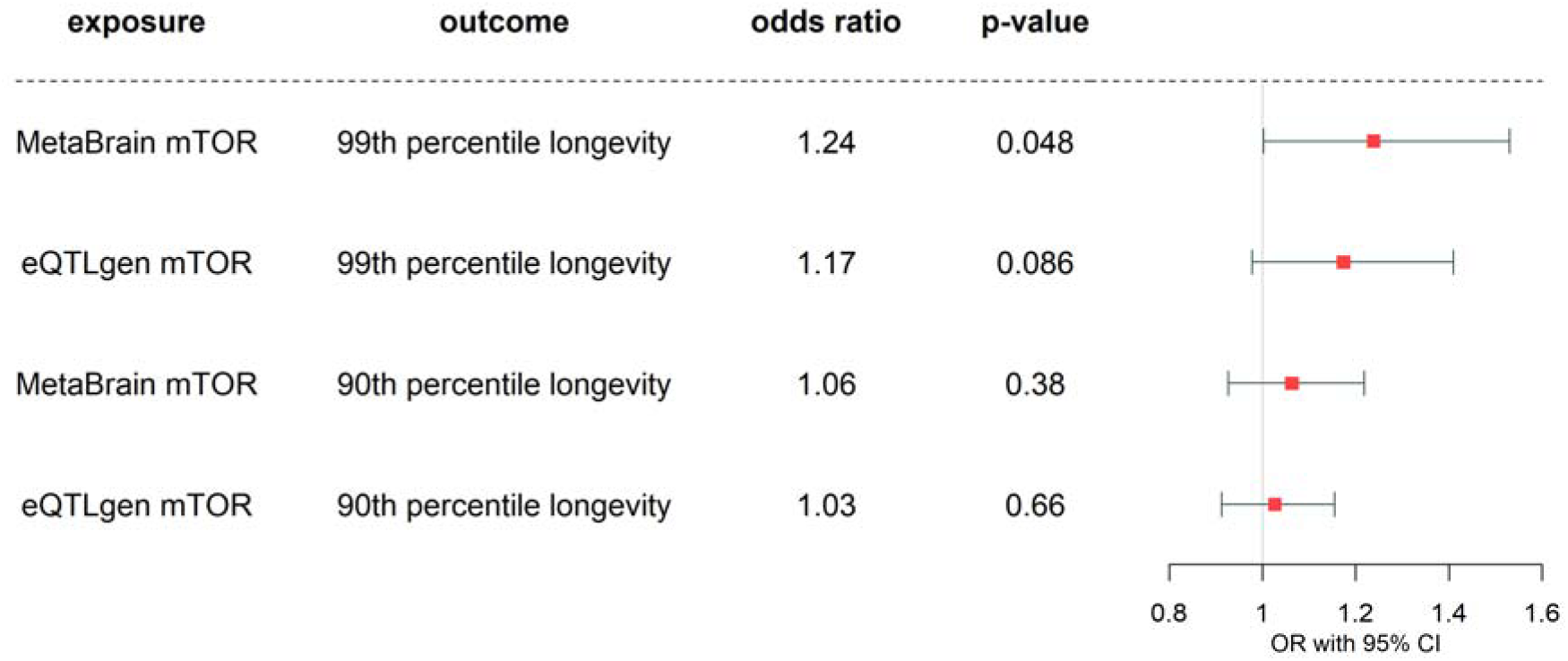
95% confidence interval of the effect of mTOR expression inhibition in the blood (eQTLgen) and in the brain cortex (MetaBrain) on the odds of belonging to the top 90^th^ or 99^th^ percentile with highest longevity. Effect sizes are scaled per 1-SD exchange in expression.

### mTOR expression affects metabolism rate and risk of type 2 diabetes along with other anthropometric traits

We then assessed the effect of *mTOR* gene expression inhibition on other outcomes, which include anthropometric traits (**Figure 3**) and potential off-target effects of rapamycin previously described in the literature (**Figure 4**). Since consistent results were obtained using both eQTLgen and MetaBrain instruments, we will focus on the estimates from eQTLgen in the following section. We found strong evidence that genetically proxied mTOR inhibition leads to reduced body mass index (β=-0.038, β_95%CI_=(−0.048,-0.028), p-value=9.4 x 10^-12^), height (β=-0.056, β _95%CI_=(−0.074,-0.038), p-value=1 x 10^-8^), age at menopause (β=-0.032, β _95%CI_=(−0.055,-0.008), p-value=0.021) but also basal metabolic rate (β=-0.04, β _95%CI_=(−0.048,-0.032), p-value=1.6 x 10^-20^). We also found evidence that bone mineral density increased on mTOR inhibition: total (β=0.099, β _95%CI_=(0.032,0.166), p-value=0.014) and heel (β=0.056, β _95%CI_=(0.034,0.078), p-value=6.4 x 10^-7^). Strong colocalisation evidence (posterior probability of H4 ranging from 0.63 to 0.95) was found for body mass index, height, heel bone mineral density, and basal metabolic rate (but not in eQTLgen).

**Figure 3.**
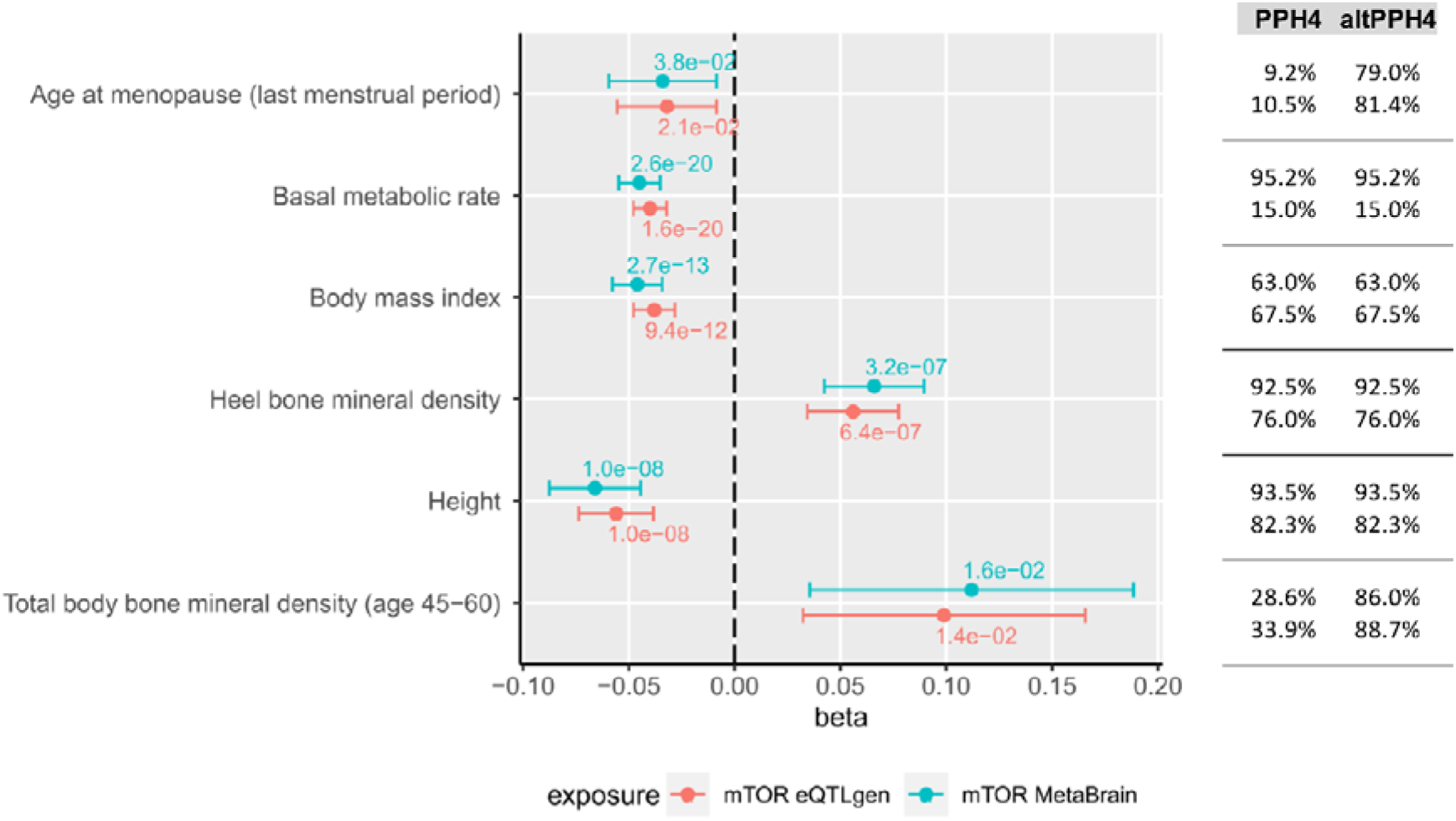
The effect of mTOR expression inhibition in the blood (eQTLgen) and in the brain cortex (MetaBrain) on a range on anthropometric outcomes. Effect sizes are scaled per 1-SD exchange in expression. P-values following FDR (Benjamini-Hochberg) correction are displayed above the 95% confidence intervals of the point estimate. Posterior probability of H4 (PPH4, true colocalisation) and alternative posterior probability of H4 colocalisation values (altPPH4) are also listed in the accompanying table.

**Figure 4.**
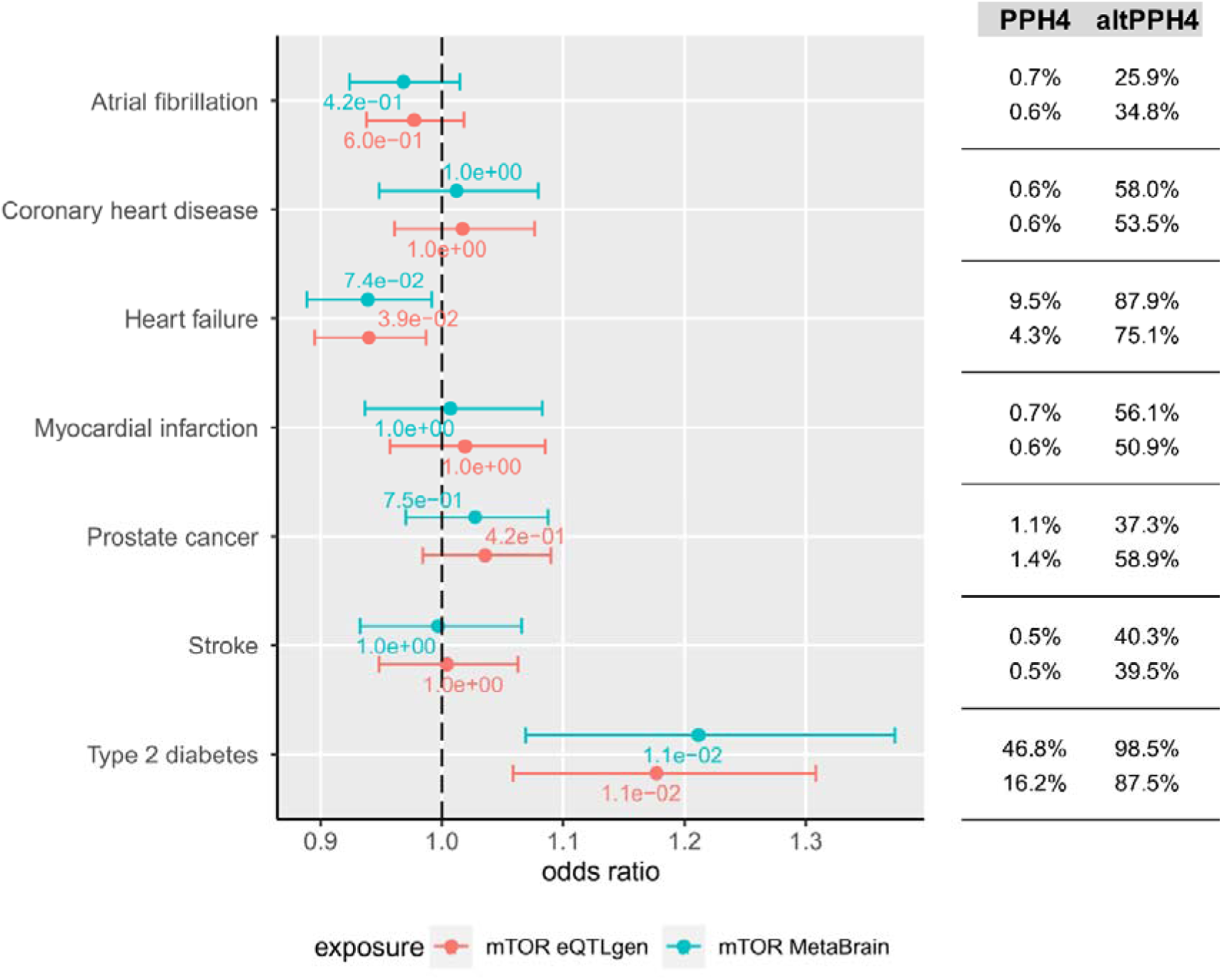
The effect of mTOR expression inhibition in the blood (eQTLgen) and in the brain cortex (MetaBrain) on type 2 diabetes, prostate cancer and cardiovascular outcomes. Effect sizes are scaled per 1-SD exchange in expression. P-values following FDR (Benjamini-Hochberg) correction are displayed above the 95% confidence intervals of the point estimate. Posterior probability of H4 (PPH4, true colocalisation) and alternative posterior probability of H4 colocalisation values (altPPH4) are also listed in the accompanying table.

There is poor MR evidence for the role of *mTOR* expression inhibition in cardiovascular disease incidence (**Figure 4**). We arrived at null results for atrial fibrillation, coronary heart disease, myocardial infarction and stroke (as well as cardiac function traits, Supplementary Table 9) but some protective effect was found for heart failure (OR=0.94, OR_955CI_=0.89-0.99, p-value=0.039) with supporting altH4 colocalisation evidence (0.75). Similarly, we found little evidence that chronic mTOR inhibition causally affects prostate cancer incidence (OR=1.04, OR_955CI_=0.98-1.09, p-value=0.423). However, we find some evidence that mTOR inhibition results in increased odds of developing type 2 diabetes (OR=1.18, OR_955CI_=1.06-1.31, p-value=0.011) which is supported by altH4 colocalisation (0.87).

### Ras-related GTP binding A expression is potentially associated with increased longevity

Having focussed on proxying the inhibition of *mTOR* by rapamycin in the first instance, we then extended our analysis to members of the mTOR complex 1 and 2, as well as proteins directly upstream and downstream of mTOR (total *n*=47) to evaluate their independent causal relationships with the select outcomes (**Supplementary Table 10**). We discovered limited, and partially contradictory, genetic support for direct effects of other mTOR pathway members on longevity (**Figure 5**, **Supplementary Figure 1**) for 6 genes, however only 1 result survived false discovery rate (FDR) correction for multiple testing. Increased expression of *RRAGA* in blood is associated with reduced odds of top 1% longevity (OR=0.41, OR_955CI_=0.24-0.72, p-value=0.035), and this result is corroborated by colocalisation (altH4=0.94). However, the effect is not replicated using MetaBrain eQTL as instruments (OR=1.31, OR_955CI_=0.81-2.12, p-value=0.65).

**Figure 5.**
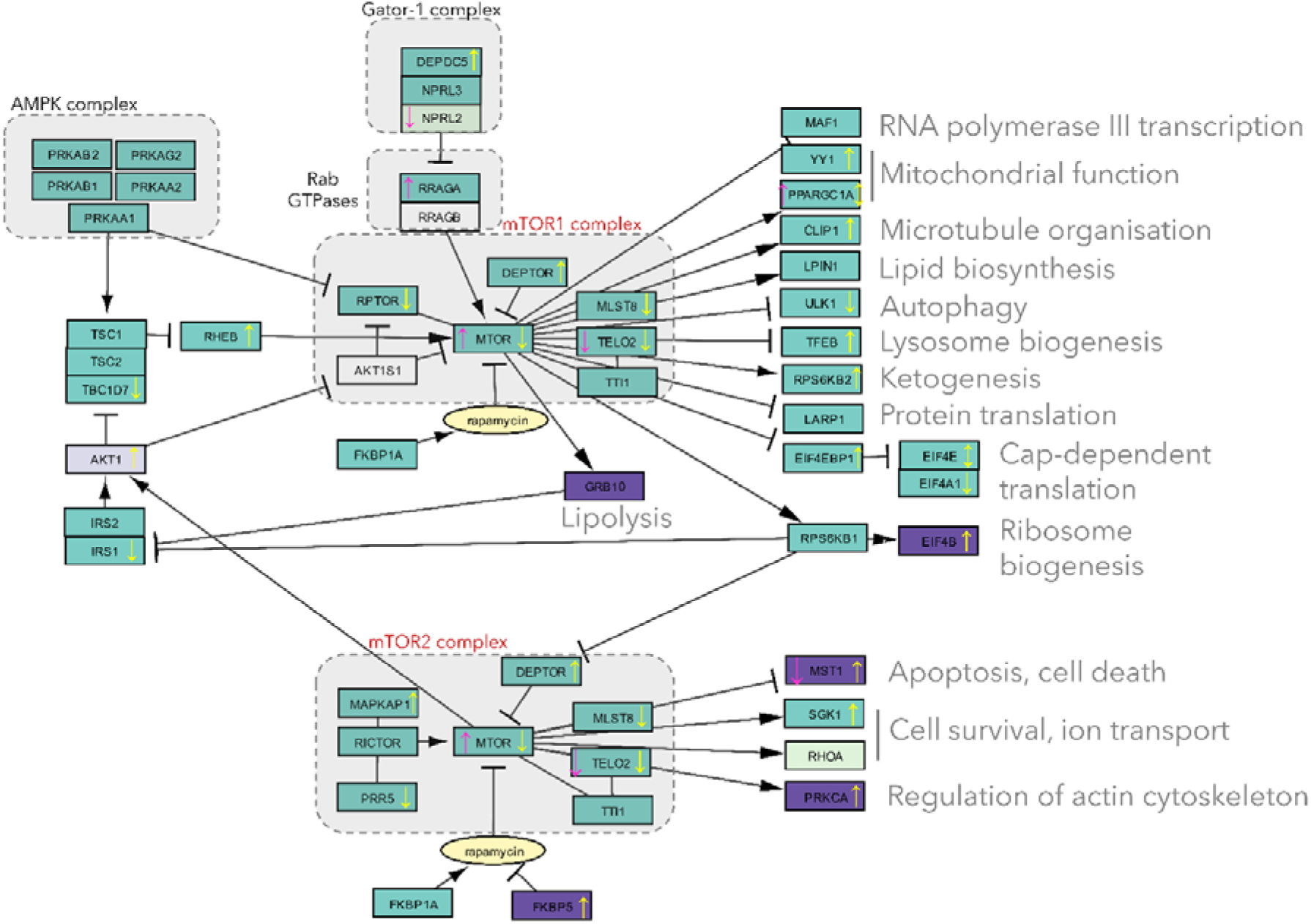
A cartoon representing mTOR1 and mTOR2 complexes and simplified network of their interactions with nutrient sensors and downstream effectors regulating metabolic processes. Each rectangle represents an individual gene product, and is coloured according to the availability of molecular QTL instruments for Mendelian Randomization. Dark teal corresponds to presence of eQTL instruments below the stringent p-value threshold of 5 x 10^-8^, whereas light teal corresponds to relaxed p-value threshold of 5 x 10^-6^. Similarly, dark purple corresponds to presence of pQTL instruments below the stringent p-value threshold (5 x 10^-8^) in addition to eQTL, whereas light purple corresponds to relaxed p-value threshold of 5 x 10^-6^. Coloured arrows within boxes represent the nominally significant (p-value < 0.05) predicted genetic effect of expression inhibition of a given gene on basal metabolic rate (yellow) and longevity (pink).

### MR evidence for effect of EIF4B and TELO2 expression on prostate cancer risk, as well as RPTOR and MAPKAP1 on coronary heart disease risk

Among disease outcomes, we found substantial evidence for the effect of mTORC pathway proteins on prostate cancer and coronary heart disease (**Supplementary Table 10**). Genetically increased protein expression of EIF4B leads to reduced risk of prostate cancer in the pQTL MR with OR=0.63 (ARIC: OR_95%CI_=0.52-0.76, p-value=1.3 x 10^-4^) with eQTL MR results in agreement. Some evidence also points to reduced gene expression of *TELO2*, member of both mTOR complexes, being protective against prostate cancer (eQTLgen: OR=0.9, OR_95%CI_=0.84-0.96, p-value=0.03). Reduced gene expression of *RPTOR* and *MAPKAP1*, components of mTOR complex 1 and 2, respectively are also potentially causally associated with reduced risk of coronary heart disease (eQTLgen *RPTOR* OR=0.91, OR_95%CI_=0.87-0.96, p-value= 0.01; eQTL *MAPKAP1* OR=0.83, OR_95%CI_=0.74-0.93, p-value=0.03).

### Members of the mTOR 1 and 2 complex pathway are overrepresented for causal impact on metabolism and BMI

Overall, across all outcomes, the highest number of genes was found to be nominally causally associated with basal metabolic rate (*n*=25, **Supplementary Figure 2**), followed by BMI (*n*=20, **Supplementary Figure 3**), and then height (*n*=16). Since as many as 54% and 29% of the mTOR pathway genes were revealed to have a nominally and FDR-corrected significant causal impact on BMR, respectively (**Figure 5**), we decided to investigate if this proportion represents enrichment relative to background selection of random genes with a genetic instrument in the eQTLgen study. We detected an overrepresentation of genes associated with BMR in the entire mTOR dataset (p-value=0.05, **Supplementary Figure 4 A**), but not H4 or altH4 coloc evidence (p-value=0.43, **Supplementary Figure 5**). We were also interested to see if the molecular components of the mTORC1 show overrepresentation as 5 out of 7 of them were found to be nominally associated with BMR: *mTOR*, Raptor, Deptor, *AKT1S1* (proxied by *AKT1* due to lack of genetic instrument) and *MLST8*; similarly for mTORC2 genes where association was found for 6 out of 8 genes (**Figure 5**). These complexes showed enrichment consistent with the extended selection of mTOR pathway genes (p-value=0.05, **Supplementary Figure 4 B**). Assessment of targets influencing body mass index revealed lack of substantial enrichment in the extended mTOR pathway gene set (p-value=0.10, **Supplementary Figure 6 A**, **Supplementary Figure 7**) but just among the 7 mTORC1 complex members (p-value=0.03, **Supplementary Figure 6 B**).

## Discussion

In this study, we used MR to evaluate whether inhibition of mammalian target of rapamycin (as proxied by eQTL in the blood and brain) causally affects longevity as well as other traits, including disease. Such a study design aimed to mimic chronic rapamycin usage and foreshadow the results of clinical trials. We find some limited evidence to support the hypothesis that inhibition of mTOR results in increased odds of reaching the top 1% of longest lifespans, possibly mediated by the effect on basal metabolic rate. Potential off-target effects include increased risk of type 2 diabetes but also reduction in the risk of heart failure. We further assessed the influence of mTOR inhibition on other traits, discovering that it substantially decreases body mass index, basal metabolic rate and height but increases Broadening our focus to encompass members of the mTOR complex 1 and 2 pathways and associated proteins, we discovered a pronounced overrepresentation of genes impacting BMR within the entire mTOR dataset, and a similar enrichment for BMI and BMR was observed for specific mTOR complexes. bone mineral density. We then extended exposure selection to include both mTORC1 and mTORC2 complexes along with their upstream regulators and downstream targets. We find enrichment of mTOR pathway genes whose expression impacts significantly on BMR relative to background of random genes with eQTL in the blood, and some evidence of their causal role in prostate cancer and coronary heart disease.

Previous MR research has fruitfully harnessed pQTL and eQTL to mimic mechanism of action of prospective or existing therapeutics^65^. Across many disease traits, association with eQTL instruments (which we focus on in our analysis of mTOR pathway) in MR revealed 2.7x enrichment of existing drug targets^66^, demonstrating the predictive power of this approach. The limitations include the inability to model molecule-specific effects as MR is applied to drug targets rather than compounds^24^ so our results inform broadly about effects of rapamycin along with rapalogs. Post-transcriptional and post-translational effects are also not easily discernible with the use of instruments derived from simple measurements of mRNA levels and protein abundance. This is a serious impediment to modelling the effects of rapamycin inhibition in the mTOR pathway where phosphorylation-based activation and inhibition play important roles^67,68^. Selection of the most relevant tissue is also important in sourcing the expression genetic instruments. For that reason, we chose morphologically and functionally distinct tissues: brain and blood, but they provide consistent support for our main findings. Ideally, eQTL and pQTL results should be compared side-by-side as the direction of effect reported can differ in up to a third of cases^69^. We were only able to triangulate MR evidence derived from eQTL and pQTL instruments in a limited number of cases – e.g. the consistently protective effect of eukaryotic translation initiation factor 4B against prostate cancer.

In general, careful consideration and validation of three assumptions is necessary to ensure validity of all MR analyses^70^. The *relevance* assumption, that genetic variants used as instrumental variables are strongly associated with the exposure of interest^22^, was satisfied by the use of strong instruments (mean F-statistics=539) associated with exposure at genome-wide significant p-value threshold of 5 x 10^-8^ (5 x 10^-6^ in a limited number of cases) and explaining a large percentage of variance in gene expression (mean=4.3%). The second assumption of *exchangeability* (association between genetic instrument and outcome is not affected by any unmeasured confounders) was evaluated by conducting colocalization comparing eQTL/pQTL signal with outcome GWAS. The exclusion restriction assumption of no horizontal pleiotropy, ie. no other independent pathways through which the exposure variant(s) can affect the outcome, cannot be thoroughly tested in our dataset due to use of single SNPs as instrumental variables^71^. However, we note that the chosen mTOR exposure SNPs show strongest evidence of association with the gene’s expression in a phenome-wide association study (PheWAS) search on OpenGWAS, and we only used *cis-* variants located in close proximity to the target gene, which should limit the risk of bias^58^.

Studies in model organisms indicate that the timing of administration of rapamycin for longevity-enhancing benefit may be crucial with effects dependent on intervention start and duration^72^. However, our MR estimates represent life-long consequences of perturbing a drug target and so they may not depict the magnitude of a drug intervention’s effect but rather inform us about its presence and direction^73^. Furthermore, MR estimates cannot easily distinguish life-time effect from effects obtainable only during critical times of development, unless multiple exposure measurements are taken across the lifecourse^74^.

Overall, we find that putatively causal relationships between mTOR and anthropometric or metabolic outcomes detected by MR triangulate well with external evidence. We find that mTOR inhibition (as proxied by lower gene expression of the mTOR kinase) results in both reduced basal metabolic rate and body mass index, and increased odds of type 2 diabetes. All of these traits have been found to have an inverse association with lifespan in other MR studies^75,76^. One caveat of MR analyses, though, is that BMR was not measured directly in the UK Biobank but estimated based on BMI, age, sex and height so the effect sizes will be correlated to those of composite traits^77^.

Reduction in BMR predisposing to longevity is in agreement with the “rate-of-living” theory of aging which links higher metabolic rate with oxidative stress, and is supported by RCT evidence in humans^78^. In animal models, mTOR inhibition by rapamycin causes reduction in BMR^50^, concomitant with increased longevity. In longitudinal human cohort studies, higher BMR has been identified as a risk factor for mortality^79,80^. In rats and mice, chronic rapamycin treatment has been reported to reduce body weight, adiposity and fat cell number whilst promoting insulin resistance and diabetes-like syndrome^81–84^. However, our results disagree with evidence from mice in which insulin sensitivity is reversed after even longer exposure to rapamycin^85^. Diabetogenic properties of rapamycin and rapalogs have long been observed in transplant patients (adjusted hazard ratio of 1.6)^19^ and are supported by adverse effect monitoring in RCTs^86,87^.

Lower height being causally associated with mTOR inhibition in our study agrees with evidence from paediatric renal transplant patients in whom rapamycin treatment for 12 months was correlated with decelerated growth rate in a case-control study setting^88^. Similarly, data from breast cancer patients are consistent with improved bone mass following rapamycin treatment since they indicate reduced bone turn-over^89,90^. Unlike other immunosuppressants, rapamycin use in rats was protective against bone loss, mainly through inhibition of bone resorption^91^.

One counterintuitive MR result obtained by us is a reduction in age at menopause caused by genetically-proxied mTOR inhibition. Rapamycin is currently being evaluated for a favourable effect on reproductive aging in humans in the VIBRANT trial (ClinicalTrials.Gov NCT05836025) since it was found to increase ovarian lifespan in mice previously^92,93^. Furthermore, in a prospective cohort study, women with early onset of menopause had a shorter life expectancy (−3.3 years) relative to healthy women experiencing menopause at a normal age^94^.

A key novelty of our study is that we deliver a bird-eye’s view of the effect of gene expression change across key players in a well-characterised biological pathway, covering both upstream and downstream targets of mTOR kinase. Alignment in the direction and size of causal effect in downstream signalling cascade could in principle be used as a robustness check against horizontal pleiotropy in our main mTOR results. However, with a complex network of protein interactions arranged into regulatory feedback loops^95^ complicating detailed quantitative interpretation, we instead focussed on global evidence of enrichment of metabolic outcomes: BMI and BMR, helping illuminate the mTOR pathway’s contribution to polygenic traits.

Rapamycin and mTOR pathway are potentially interesting targets for promoting life extension in humans, however the risk of side effects (such as T2D) requires caution in any RCT with rapamycin and its analogs used as intervention. Based on MR evidence, well-informed off-label chronic use of rapamycin by healthy “biohackers” needs to balance the potential cost of decreased healthspan relative to benefit of increased longevity. Future MR studies of the mTOR pathway could be improved by better powered source GWAS and generation of pQTL resources for more gene targets.

## Data and code availability

GWAS summary statistics availability is provided by relevant publications. We accessed the following summary statistics via OpenGWAS: ebi-a-GCST005350, ebi-a-GCST006288, ebi-a-GCST006414, ebi-a-GCST006906, ebi-a-GCST009541, ieu-a-7, ieu-a-26, ieu-a-89, ieu-a-798, ieu-b-40, ieu-b-85, ukb-b-17422, ukb-b-16446.

## Author contributions

MKS: Conceptualization, Methodology, Formal analysis, Visualization, Writing – original draft, Writing – review and editing

TG: Supervision, Methodology, Resources, Writing – review and editing, Funding acquisition

---

**Supplementary Figure 1.**
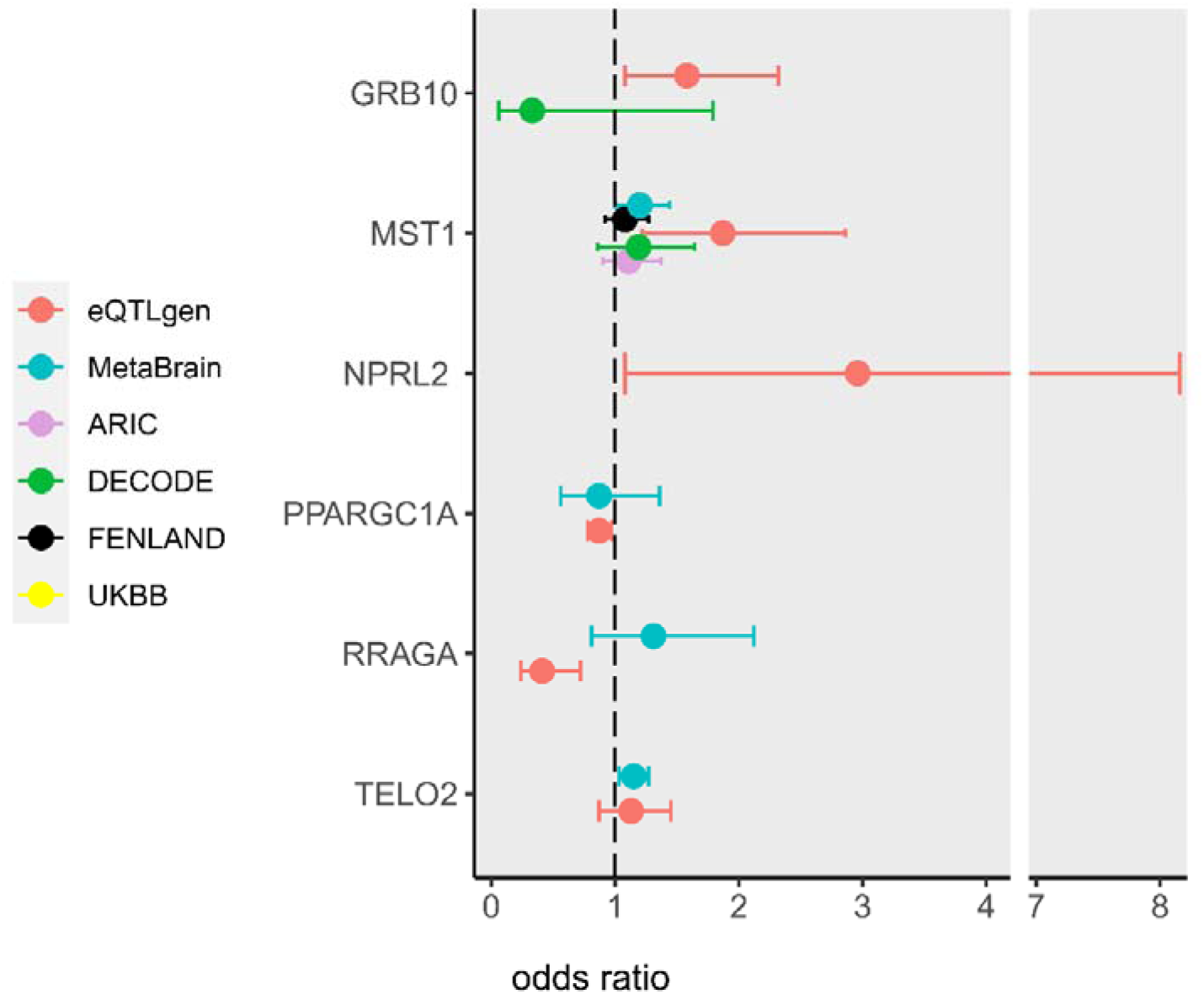
95% confidence intervals of the effect of gene expression of nominally significant mTOR pathway genes in the blood (eQTL: eQTLgen, pQTL: ARIC, deCODE, FENLAND, UKBB) and brain cortex (eQTL: MetaBrain) on the odds of belonging to the 99^th^ percentile with highest longevity. Effect sizes are scaled per 1-SD exchange in expression.

**Supplementary Figure 2.**
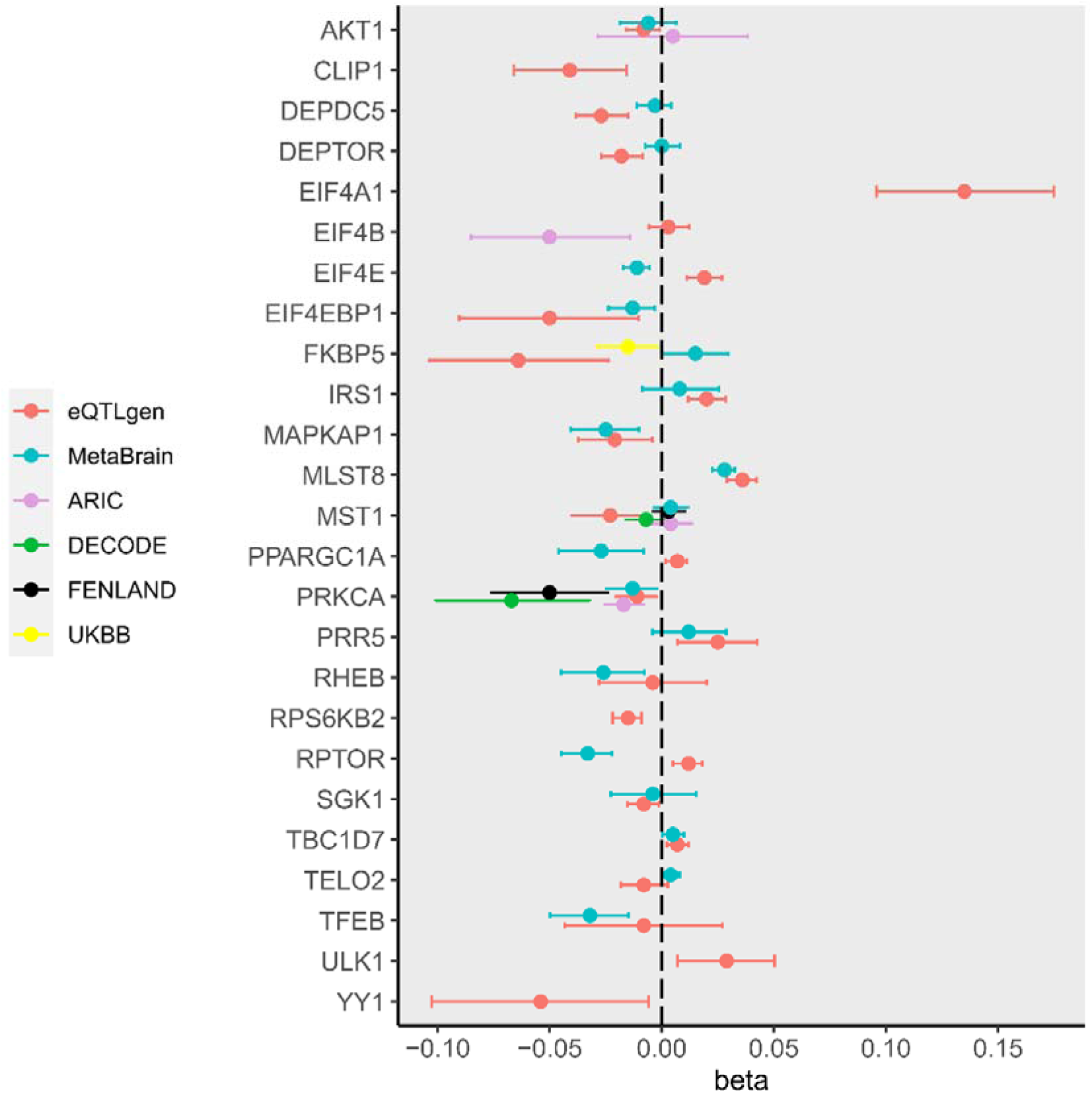
95% confidence intervals of the effect of gene expression of nominally significant mTOR pathway genes in the blood (eQTL: eQTLgen, pQTL: ARIC, deCODE, FENLAND, UKBB) and brain cortex (eQTL: MetaBrain) on basal metabolic rate (BMR). Effect sizes are scaled per 1-SD exchange in expression.

**Supplementary Figure 3.**
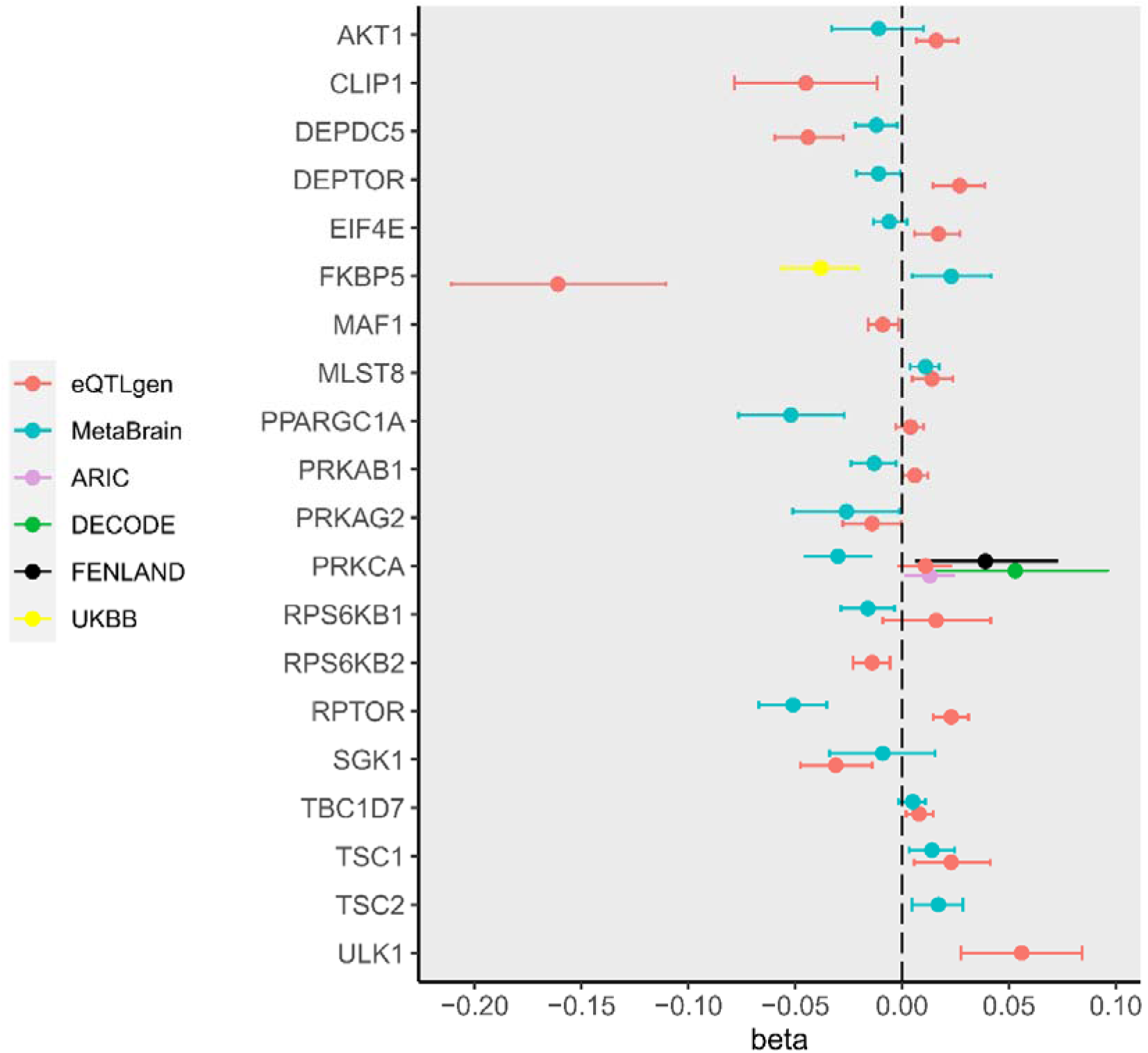
95% confidence intervals of the effect of gene expression of nominally significant mTOR pathway genes in the blood (eQTL: eQTLgen, pQTL: ARIC, deCODE, FENLAND, UKBB) and brain cortex (eQTL: MetaBrain) on body mass index (BMI). Effect sizes are scaled per 1-SD exchange in expression.

**Supplementary Figure 4.**
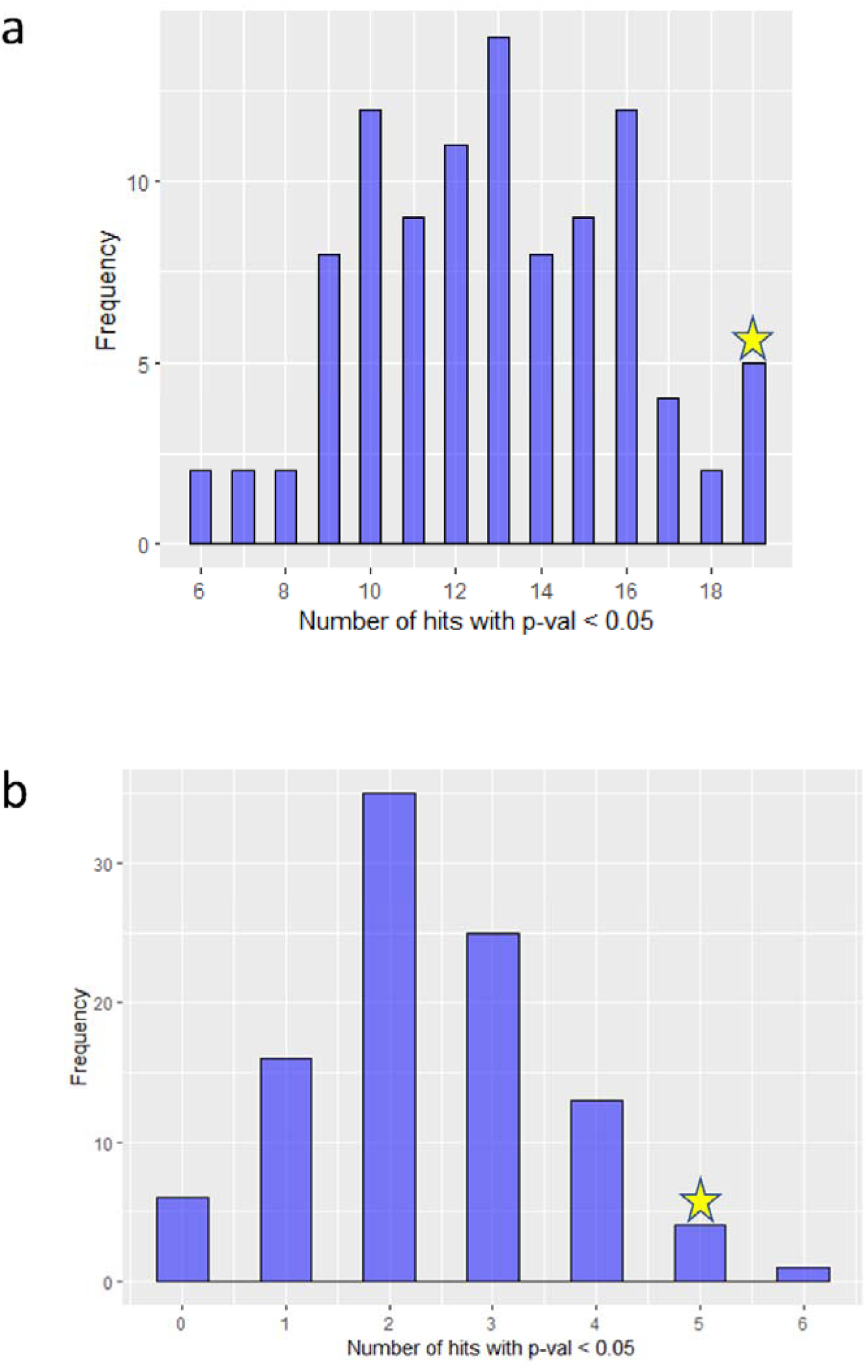
Distribution of significant (nominal p-value < 0.05) MR results among 100 replicates of a set of randomly selected genetic instruments from eQTLgen blood eQTL dataset with basal metabolic rate (BMR) used as outcome: a) set of 48 genetic instruments; b) set of 7 genetic instruments. The result observed for mTOR pathway gene set is highlighted with a golden star.

**Supplementary Figure 5.**
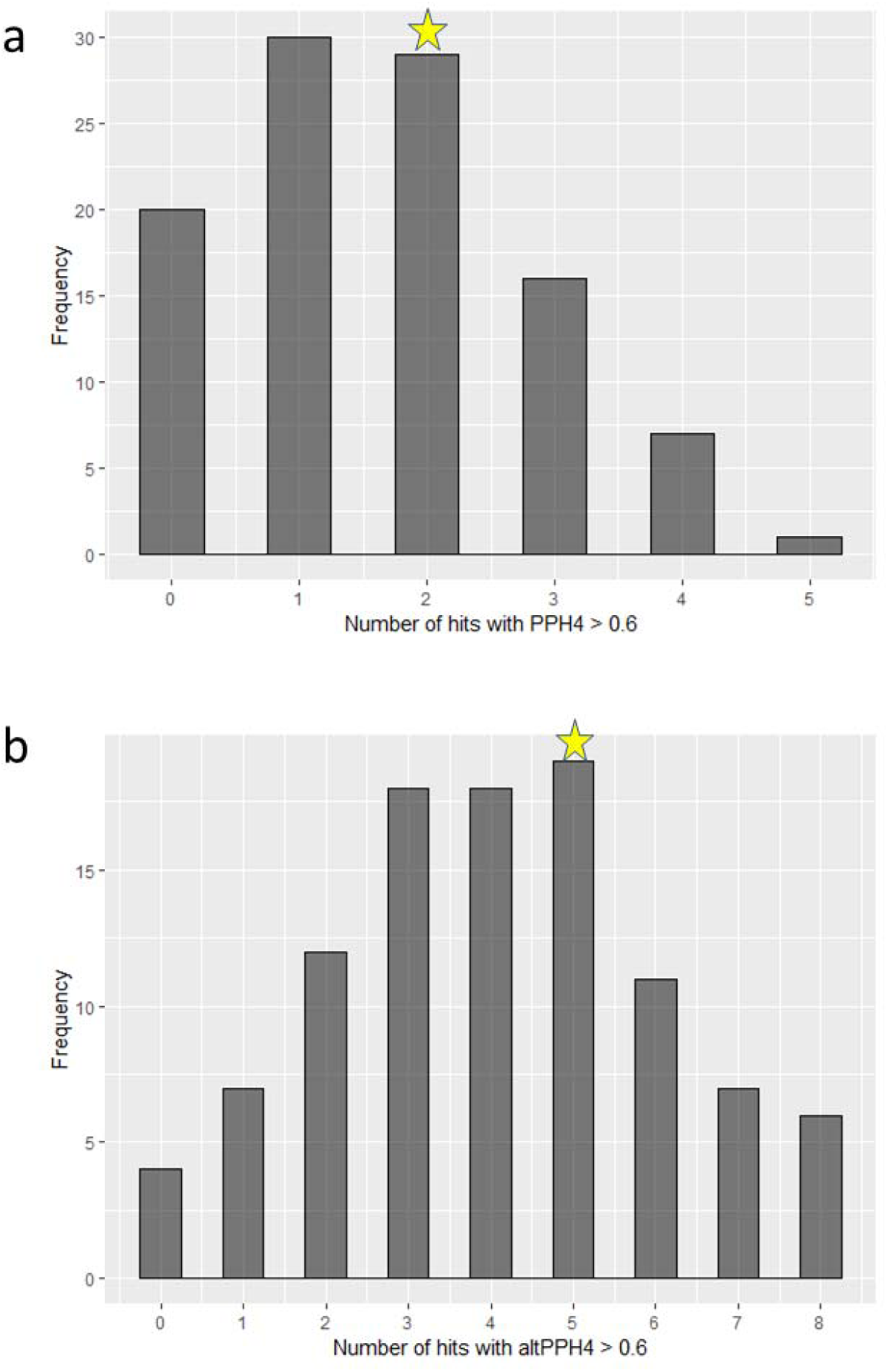
Distribution of coloc results among 100 replicates of randomly selected 48 genetic instruments from eQTLgen blood eQTL dataset with basal metabolic rate (BMR) used as outcome. Coloc was applied only to nominally significant MR results. a) number of coloc results with posterior probability (PP) of hypothesis 4 (H4) > 0.6; b) number of coloc results with alternative posterior probability (PP) of hypothesis 4 (H4) > 0.6.

**Supplementary Figure 6.**
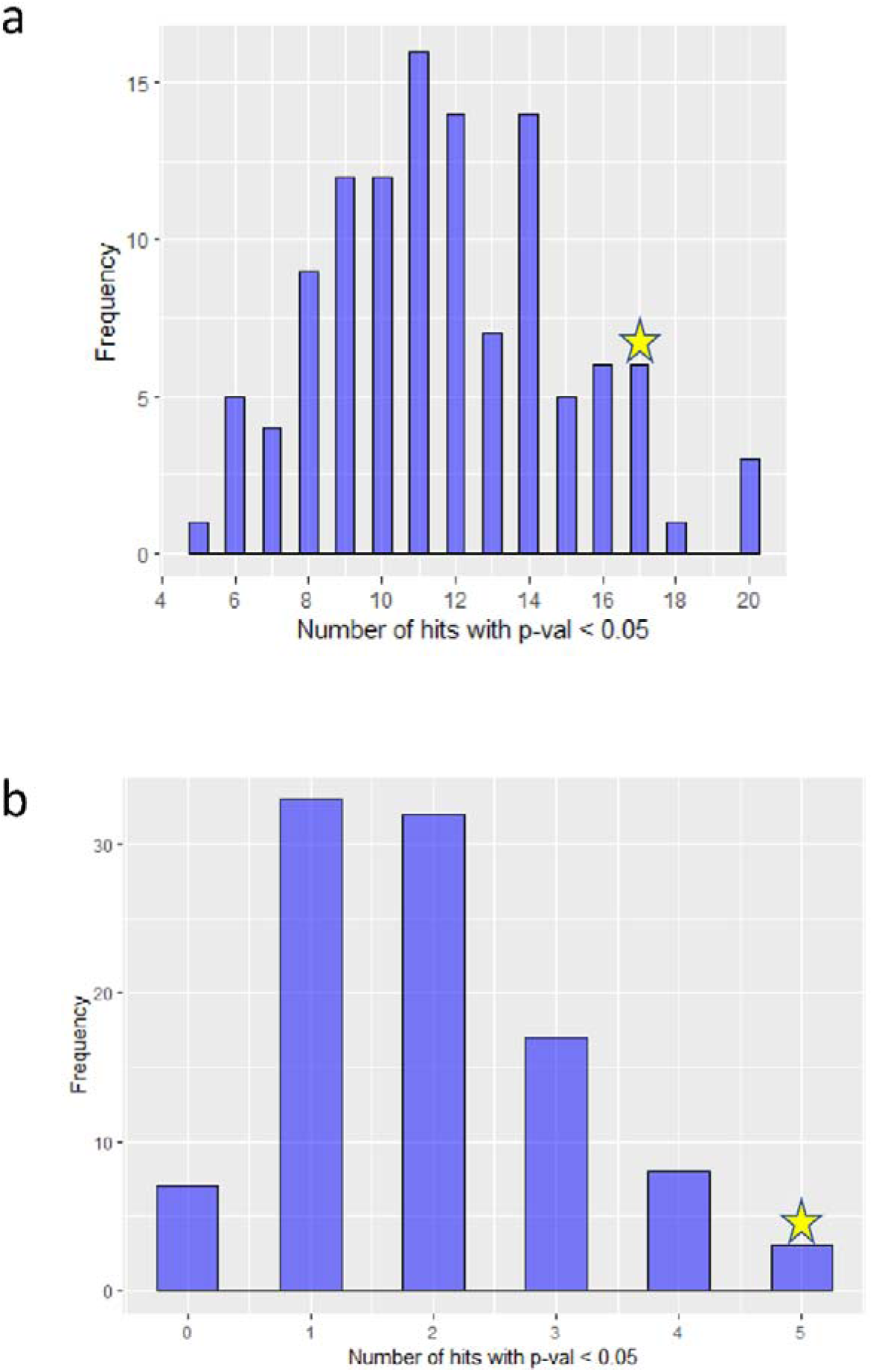
Distribution of significant (nominal p-value < 0.05) MR results among 100 replicates of a set of randomly selected genetic instruments from eQTLgen blood eQTL dataset with body mass index (BMI) used as outcome: a) set of 48 genetic instruments; b) set of 7 genetic instruments. The result observed for MTOR pathway gene set is highlighted with a golden star.

**Supplementary Figure 7.**
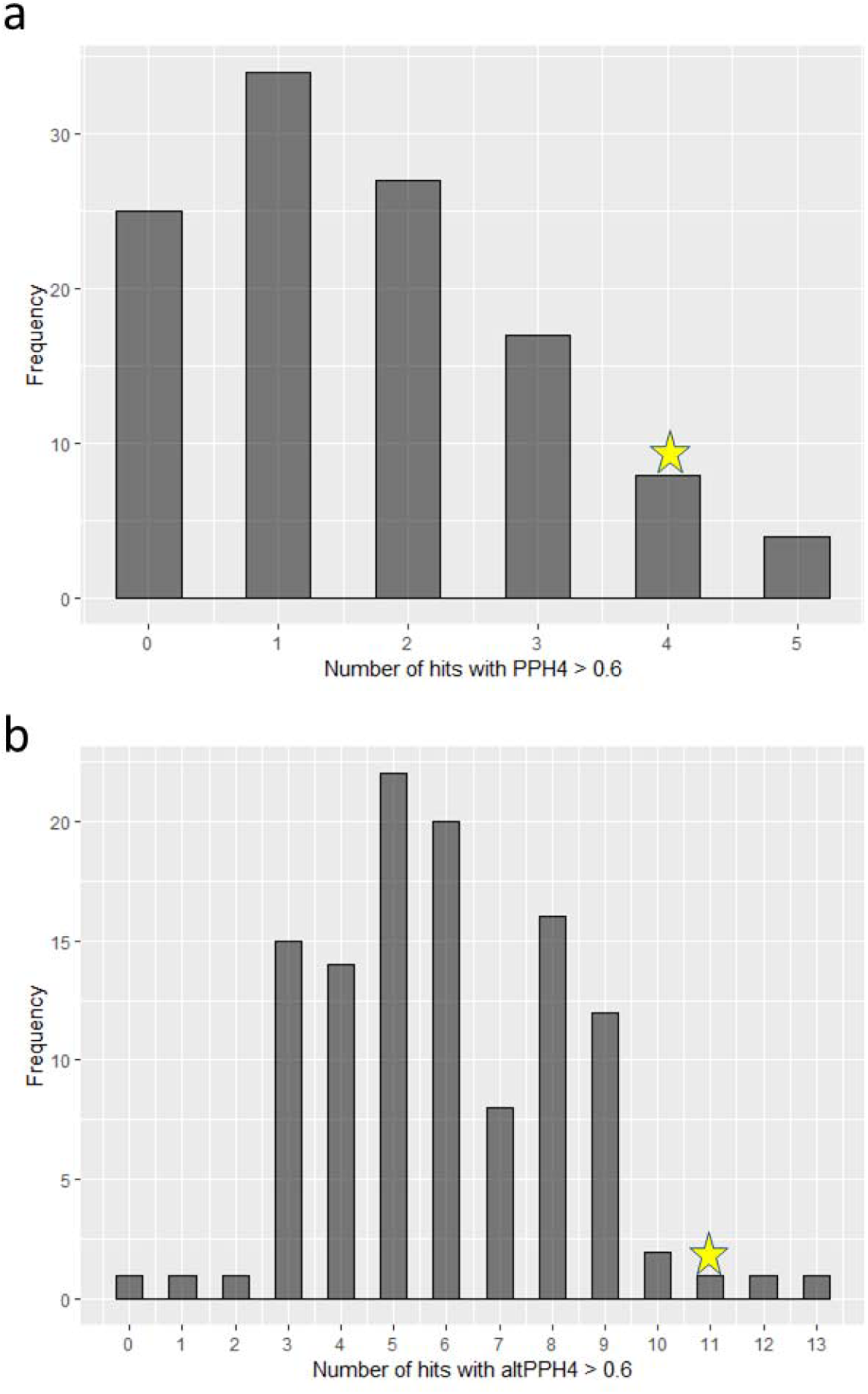
Distribution of coloc results among 100 replicates of randomly selected 48 genetic instruments from eQTLgen blood eQTL dataset with body mass index (BMI) used as outcome. Coloc was applied only to significant MR results. a) number of coloc results with posterior probability (PP) of hypothesis 4 (H4) > 0.6; b) number of coloc results with alternative posterior probability (PP) of hypothesis 4 (H4) > 0.6.

## Reference

1. Zoncu, R., Efeyan, A. & Sabatini, D. M. mTOR: from growth signal integration to cancer, diabetes and ageing. Nat. Rev. Mol. Cell Biol. 12, 21–35 (2011).

2. Simcox, J. & Lamming, D. W. The central moTOR of metabolism. Dev. Cell 57, 691–706 (2022).

3. Papadopoli, D. et al. Mtor as a central regulator of lifespan and aging. F1000Research 8, (2019).

4. Kennedy, B. K. & Lamming, D. W. The Mechanistic Target of Rapamycin: The Grand ConducTOR of Metabolism and Aging. Cell Metab. 23, 990–1003 (2016).

5. Hobby, G., Clark, R. & Woywodt, A. A treasure from a barren island: the discovery of rapamycin. Clin. Kidney J. 15, 1971–1972 (2022).

6. Lamming, D. W., Ye, L., Sabatini, D. M. & Baur, J. A. Rapalogs and mTOR inhibitors as anti-aging therapeutics. J. Clin. Invest. 123, 980–989 (2013).

7. Hartford, C. M. & Ratain, M. J. Rapamycin: Something Old, Something New, Sometimes Borrowed and Now Renewed. Clin. Pharmacol. Ther. 82, 381–388 (2007).

8. Partridge, L., Fuentealba, M. & Kennedy, B. K. The quest to slow ageing through drug discovery. Nat. Rev. Drug Discov. 19, 513–532 (2020).

9. Lamming, D. W. et al. Rapamycin-Induced Insulin Resistance Is Mediated by mTORC2 Loss and Uncoupled from Longevity. Science (80-. ). 335, 1638–1643 (2012).

10. Schreiber, K. H. et al. A novel rapamycin analog is highly selective for mTORC1 in vivo. Nat. Commun. 10, 3194 (2019).

11. Green, C. L., Lamming, D. W. & Fontana, L. Molecular mechanisms of dietary restriction promoting health and longevity. Nat. Rev. Mol. Cell Biol. 23, 56–73 (2022).

12. Regaldo, A. Is This the Anti-Aging Pill We’ve All Been Waiting For? MIT Technol. Rev. (2017).

13. Easter, M. This Obscure, Potentially Dangerous Drug Could Stop Aging. Men’s Heal. (2019).

14. Hamzelou, J. These scientists are working to extend the life span of pet dogs—and their owners. MIT Technol. Rev. (2022).

15. Brueck, H. Scientists and biohackers are popping this $1 cancer drug to stop their cells from aging. Insider (2023).

16. Mannick, J. B. et al. TORC1 inhibition enhances immune function and reduces infections in the elderly. Sci. Transl. Med. 10, eaaq1564 (2018).

17. Mannick, J. B. et al. Targeting the biology of ageing with mTOR inhibitors to improve immune function in older adults: phase 2b and phase 3 randomised trials. *lancet*. Heal. Longev. 2, e250–e262 (2021).

18. Garay, R. P. Investigational drugs and nutrients for human longevity. Recent clinical trials registered in ClinicalTrials.gov and clinicaltrialsregister.eu. Expert Opin. Investig. Drugs 30, 749–758 (2021).

19. Johnston, O., Rose, C. L., Webster, A. C. & Gill, J. S. Sirolimus Is Associated with New-Onset Diabetes in Kidney Transplant Recipients. J. Am. Soc. Nephrol. 19, (2008).

20. Yanik, E. L., Siddiqui, K. & Engels, E. A. Sirolimus effects on cancer incidence after kidney transplantation: a meta-analysis. Cancer Med. 4, 1448–1459 (2015).

21. Davies, N. M., Holmes, M. V. & Davey Smith, G. Reading Mendelian randomisation studies: A guide, glossary, and checklist for clinicians. BMJ 362, (2018).

22. Sanderson, E. et al. Mendelian randomization. Nat. Rev. Methods Prim. 2, 6 (2022).

23. Sobczyk, M. K., Zhang, J., Davey Smith, G. & Gaunt, T. R. Systematic comparison of Mendelian randomization studies and randomized controlled trials using electronic databases. medRxiv https://www.medrxiv.org/content/10.1101/2022.04.11.22273633v2 (2022) 10.1101/2022.04.11.22273633.

24. Gill, D. et al. Mendelian randomization for studying the effects of perturbing drug targets. Wellcome open Res. 6, 16 (2021).

25. Lin, J., Zhou, J. & Xu, Y. Potential drug targets for multiple sclerosis identified through Mendelian randomization analysis. Brain awad070 (2023) doi:10.1093/brain/awad070.

26. Chen, Y. et al. Genetic insights into therapeutic targets for aortic aneurysms: A Mendelian randomization study. eBioMedicine 83, (2022).

27. Zhao, H. et al. Proteome-wide Mendelian randomization in global biobank meta-analysis reveals multi-ancestry drug targets for common diseases. Cell Genomics 2, (2022).

28. Richardson, T. G., Hemani, G., Gaunt, T. R., Relton, C. L. & Davey Smith, G. A transcriptome-wide Mendelian randomization study to uncover tissue-dependent regulatory mechanisms across the human phenome. Nat. Commun. 11, 185 (2020).

29. Jiang, J.-C., Hu, C., McIntosh, A. M. & Shah, S. Investigating the potential anti-depressive mechanisms of statins: a transcriptomic and Mendelian randomization analysis. Transl. Psychiatry 13, 110 (2023).

30. Zheng, J. et al. Evaluating the efficacy and mechanism of metformin targets on reducing Alzheimer’s disease risk in the general population: a Mendelian randomisation study. Diabetologia 65, 1664–1675 (2022).

31. Storm, C. S. et al. Finding genetically-supported drug targets for Parkinson’s disease using Mendelian randomization of the druggable genome. Nat. Commun. 12, 7342 (2021).

32. Soliman, G. A. & Schooling, C. M. Causal association between mTOR-dependent EIF-4E and EIF-4A circulating protein levels and type 2 diabetes: a Mendelian randomization study. Sci. Rep. 10, (2020).

33. Cai, Y. et al. Association of mTORC1-dependent circulating protein levels with cataract formation: a mendelian randomization study. BMC Genomics 23, (2022).

34. Mu, Y.-F. et al. Genetic Evidence Supporting Causal Roles of mTOR-Dependent Proteins in Rheumatic Fever: A Two-Sample Randomized Mendelian Study. Adv. Ther. (2023) doi:10.1007/s12325-022-02419-4.

35. Tan, C., Ai, J. & Zhu, Y. mTORC1-Dependent Protein and Parkinson’s Disease: A Mendelian Randomization Study. Brain Sci. 13, (2023).

36. Cai, H.-Y. et al. Causal Association Between mTOR-Dependent Protein Levels and Alzheimer’s Disease: A Mendelian Randomization Study. J. Alzheimer’s Dis. Preprint, 1–9 (2023).

37. Kanehisa, M., Furumichi, M., Tanabe, M., Sato, Y. & Morishima, K. KEGG: new perspectives on genomes, pathways, diseases and drugs. Nucleic Acids Res. 45, D353–D361 (2017).

38. de Klein, N. et al. Brain expression quantitative trait locus and network analyses reveal downstream effects and putative drivers for brain-related diseases. Nat. Genet. 55, 377–388 (2023).

39. Võsa, U. et al. Large-scale cis- and trans-eQTL analyses identify thousands of genetic loci and polygenic scores that regulate blood gene expression. Nat. Genet. 2021 539 53, 1300–1310 (2021).

40. Zhang, J. et al. Plasma proteome analyses in individuals of European and African ancestry identify cis-pQTLs and models for proteome-wide association studies. Nat. Genet. (2022) doi:10.1038/s41588-022-01051-w.

41. Ferkingstad, E. et al. Large-scale integration of the plasma proteome with genetics and disease. Nat. Genet. 53, 1712–1721 (2021).

42. Pietzner, M. et al. Mapping the proteo-genomic convergence of human diseases. Science (80-. ). 374, eabj1541 (2021).

43. BB, S., et al. Genetic regulation of the human plasma proteome in 54,306 UK Biobank participants. Popul. Anal. Janssen Data Sci. 20, (2022).

44. The GTEx Consortium atlas of genetic regulatory effects across human tissues. Science (80-. ). 369, 1318–1330 (2020).

45. Takei, N. & Nawa, H. mTOR signaling and its roles in normal and abnormal brain development. Frontiers in Molecular Neuroscience vol. 7 (2014).

46. Schmidt, M. et al. The Human Blood Transcriptome in a Large Population Cohort and Its Relation to Aging and Health. *Front*. Big Data 3, (2020).

47. Aguet, F. et al. Genetic effects on gene expression across human tissues. Nature 550, 204–213 (2017).

48. Deelen, J. et al. A meta-analysis of genome-wide association studies identifies multiple longevity genes. Nat. Commun. 2019 101 10, 1–14 (2019).

49. Liu, Y. et al. EpiGraphDB: a database and data mining platform for health data science. Bioinformatics 0–0 (2020).

50. Schieke, S. M. et al. The Mammalian Target of Rapamycin (mTOR) Pathway Regulates Mitochondrial Oxygen Consumption and Oxidative Capacity*. J. Biol. Chem. 281, 27643–27652 (2006).

51. Valvezan, A. J. & Manning, B. D. Molecular logic of mTORC1 signalling as a metabolic rheostat. Nat. Metab. 1, 321–333 (2019).

52. McMullen, J. R. et al. Inhibition of mTOR Signaling With Rapamycin Regresses Established Cardiac Hypertrophy Induced by Pressure Overload. Circulation 109, 3050–3055 (2004).

53. Flynn, J. M. et al. Late-life rapamycin treatment reverses age-related heart dysfunction. Aging Cell 12, 851–862 (2013).

54. Quarles, E. et al. Rapamycin persistently improves cardiac function in aged, male and female mice, even following cessation of treatment. Aging Cell 19, e13086 (2020).

55. Rai, J. S., Henley, M. J. & Ratan, H. L. Mammalian target of rapamycin: A new target in prostate cancer. Urol. Oncol. Semin. Orig. Investig. 28, 134–138 (2010).

56. Kemp Bohan, P. M., et al. Results of a phase Ib trial of encapsulated rapamycin in prostate cancer patients under active surveillance to prevent progression. J. Clin. Oncol. 38, 34 (2020).

57. Elsworth, B., et al. The MRC IEU OpenGWAS data infrastructure. bioRxiv (2020) doi:10.1101/2020.08.10.244293.

58. Holmes, M. V, Richardson, T. G., Ference, B. A., Davies, N. M. & Davey Smith, G. Integrating genomics with biomarkers and therapeutic targets to invigorate cardiovascular drug development. Nat. Rev. Cardiol. 18, 435–453 (2021).

59. McLaren, W. et al. The ensembl variant effect predictor. Genome Biol. 17, 1–14 (2016).

60. Auton, A. et al. A global reference for human genetic variation. Nature 526, 68–74 (2015).

61. Zheng, X. et al. A high-performance computing toolset for relatedness and principal component analysis of SNP data. Bioinformatics 28, 3326–3328 (2012).

62. Hemani, G. et al. The MR-Base platform supports systematic causal inference across the human phenome. Elife 7, e34408 (2018).

63. Giambartolomei, C. et al. Bayesian Test for Colocalisation between Pairs of Genetic Association Studies Using Summary Statistics. PLoS Genet. 10, (2014).

64. Zuber, V. et al. Combining evidence from Mendelian randomization and colocalization: Review and comparison of approaches. Am. J. Hum. Genet. (2022) 10.1016/j.ajhg.2022.04.001.

65. Gill, D. & Burgess, S. The evolution of mendelian randomization for investigating drug effects. PLOS Med. 19, 1–3 (2022).

66. Sadler, M. C., Auwerx, C., Deelen, P. & Kutalik, Z. Multi-layered genetic approaches to identify approved drug targets. Cell Genomics 3, 100341 (2023).

67. Kang, S. A. et al. mTORC1 Phosphorylation Sites Encode Their Sensitivity to Starvation and Rapamycin. Science (80-. ). 341, 1236566 (2013).

68. Battaglioni, S., Benjamin, D., Wälchli, M., Maier, T. & Hall, M. N. mTOR substrate phosphorylation in growth control. Cell 185, 1814–1836 (2022).

69. Robinson, J. W., et al. Evaluating the potential benefits and pitfalls of combining protein and expression quantitative trait loci in evidencing drug targets. bioRxiv (2022) doi:10.1101/2022.03.15.484248.

70. Burgess, S. & Malarstig, A. Using Mendelian randomization to assess and develop clinical interventions: limitations and benefits. J. Comp. Eff. Res. 2, 209–212 (2013).

71. Cho, Y. et al. Exploiting horizontal pleiotropy to search for causal pathways within a Mendelian randomization framework. Nat. Commun. 11, 1010 (2020).

72. Blagosklonny, M. V. Rapamycin treatment early in life reprograms aging: hyperfunction theory and clinical practice. Aging (Albany. NY). 14, 8140–8149 (2022).

73. Gill, D., Walker, V. M., Martin, R. M., Davies, N. M. & Tzoulaki, I. Comparison with randomized controlled trials as a strategy for evaluating instruments in Mendelian randomization. Int. J. Epidemiol. 49, 1404–1406 (2020).

74. Power, G. M. et al. A systematic literature review of methodological approaches, challenges, and opportunities in the application of Mendelian randomisation to lifecourse epidemiology. medRxiv (2023) doi:10.1101/2023.05.16.22283780.

75. Ng, J. C. M. & Schooling, C. M. Effect of basal metabolic rate on lifespan: a sex-specific Mendelian randomization study. Sci. Rep. 13, 7761 (2023).

76. van Oort, S., Beulens, J. W. J., van Ballegooijen, A. J., Burgess, S. & Larsson, S. C. Cardiovascular risk factors and lifestyle behaviours in relation to longevity: a Mendelian randomization study. J. Intern. Med. 289, 232–243 (2021).

77. Kloecker, D. E., Brage, S. & Wareham, N. J. Comment on Baranova, et al. Causal Associations Between Basal Metabolic Rate and COVID-19. Diabetes 2023;72:149–154. Diabetes 72, e6–e7 (2023).

78. Redman, L. M. et al. Metabolic Slowing and Reduced Oxidative Damage with Sustained Caloric Restriction Support the Rate of Living and Oxidative Damage Theories of Aging. Cell Metab. 27, 805–815.e4 (2018).

79. Ruggiero, C. et al. High Basal Metabolic Rate Is a Risk Factor for Mortality: The Baltimore Longitudinal Study of Aging. Journals Gerontol. Ser. A 63, 698–706 (2008).

80. Jumpertz, R. et al. Higher Energy Expenditure in Humans Predicts Natural Mortality. J. Clin. Endocrinol. Metab. 96, E972–E976 (2011).

81. Houde, V. P. et al. Chronic Rapamycin Treatment Causes Glucose Intolerance and Hyperlipidemia by Upregulating Hepatic Gluconeogenesis and Impairing Lipid Deposition in Adipose Tissue. Diabetes 59, 1338–1348 (2010).

82. Deblon, N. et al. Chronic mTOR inhibition by rapamycin induces muscle insulin resistance despite weight loss in rats. Br. J. Pharmacol. 165, 2325–2340 (2012).

83. Liu, Y. et al. Rapamycin-induced metabolic defects are reversible in both lean and obese mice. Aging (Albany. NY). 6, 742–754 (2014).

84. Chang, G.-R. et al. Long-term administration of rapamycin reduces adiposity, but impairs glucose tolerance in high-fat diet-fed KK/HlJ mice. Basic Clin. Pharmacol. Toxicol. 105, 188–198 (2009).

85. Fang, Y. et al. Duration of rapamycin treatment has differential effects on metabolism in mice. Cell Metab. 17, 456–462 (2013).

86. Sivendran, S. et al. Metabolic complications with the use of mTOR inhibitors for cancer therapy. Cancer Treat. Rev. 40, 190–196 (2014).

87. Vergès, B. & Cariou, B. mTOR inhibitors and diabetes. Diabetes Res. Clin. Pract. 110, 101–108 (2015).

88. González, D. et al. Growth of kidney-transplanted pediatric patients treated with sirolimus. Pediatr. Nephrol. 26, 961–966 (2011).

89. Gnant, M. et al. Effect of Everolimus on Bone Marker Levels and Progressive Disease in Bone in BOLERO-2. JNCI J. Natl. Cancer Inst. 105, 654–663 (2013).

90. Hadji, P. et al. The impact of mammalian target of rapamycin inhibition on bone health in postmenopausal women with hormone receptor-positive advanced breast cancer receiving everolimus plus exemestane in the phase IIIb 4EVER trial. J. bone Oncol. 14, 10 (2019).

91. Goodman, G. R. et al. Immunosuppressant Use Without Bone Loss—Implications for Bone Loss After Transplantation. J. Bone Miner. Res. 16, 72–78 (2001).

92. Mok-Lin, E. et al. Premature recruitment of oocyte pool and increased mTOR activity in Fmr1 knockout mice and reversal of phenotype with rapamycin. Sci. Rep. 8, 588 (2018).

93. Dou, X. et al. Short-term rapamycin treatment increases ovarian lifespan in young and middle-aged female mice. Aging Cell 16, 825–836 (2017).

94. Asllanaj, E. et al. Age at natural menopause and life expectancy with and without type 2 diabetes. Menopause 26, 387–394 (2019).

95. Efeyan, A. & Sabatini, D. M. mTOR and cancer: many loops in one pathway. Curr. Opin. Cell Biol. 22, 169–176 (2010).

